# School-Related Public Inquiries Received by the Centers for Disease Control and Prevention During the COVID-19 Pandemic, United States, 2020-2022

**DOI:** 10.1101/2025.06.16.25329578

**Authors:** Danielle Kleven, Lisa C. Barrios, Lillian Fineman, Neha Kanade Cramer, Wrishija Roy

**Author notes:** **Corresponding Author:** Danielle Kleven, MPH, Centers for Disease Control and Prevention, National Center for Emerging and Zoonotic Infectious Diseases, Division of Infectious Disease Readiness and Innovation, 1600 Clifton Rd NE, Atlanta, GA 30333, USA.

## Abstract

**Objectives:** After supporting the COVID-19 emergency response and responding to public inquiries, we sought to identify common themes to prepare for and improve future emergency response efforts. We examined how often various themes (topics) were asked of the Centers for Disease Control and Prevention (CDC) through CDC-INFO (agents who respond to public requests) about the COVID-19 pandemic and kindergarten through grade 12 (K-12) schools, whether inquiries included questions or complaints, how theme frequency changed, and how the source of CDC-INFO agent responses varied by theme.

**Methods:** We analyzed inquiries (questions or complaints) received by CDC-INFO from January 17, 2020, to November 8, 2022. We pilot-tested our protocol by coding 300 inquiries to create a code book with 11 themes (eg, quarantine and isolation, general guidance) and then tested them for interrater reliability before coding 4000 additional randomly selected inquiries (of 24 502 inquiries received). We then compiled descriptive statistics and used Chi-square goodness-of-fit tests to compare differences between frequencies of categorical variables.

**Results:** We found 2180 inquiries related to K-12 schools and COVID-19 and assigned 1 or more themes for analysis (resulting in 2439 themes). The most common theme was quarantine and isolation (39%). Across all themes, except for closures, the frequency of questions was greater than the frequency of complaints. For 5 of 11 themes (closure, general guidance, face masks, quarantine and isolation, travel), we found significant differences in inquiry frequency over time. For all themes, except for travel, we found significant differences in which sources CDC-INFO agents used to respond to inquiries.

**Conclusions:** Public health officials should be prepared to respond to school-based questions about quarantine and isolation and to address various topics as the emergency changes over time.

---

In January 2020, the Centers for Disease Control and Prevention (CDC) began responding to the 2019 novel coronavirus (later named COVID-19) pandemic.^1^ One aspect of this response was providing guidance on preventing the spread of COVID-19 in kindergarten through grade 12 (K-12) schools. CDC provided information to the public through the CDC-INFO contact center, to which anyone can call or email with a question, complaint, or comment.^2^ The term “inquiry” is an umbrella term that describes all telephone calls or emails to CDC-INFO, regardless of content. The CDC-INFO contact center is staffed by agents who receive the inquiries and respond to them. Agents use prepared responses or the CDC website to provide answers to common questions and escalate any complex inquiries to CDC experts. Agents track information in a secure system, such as the inquiry’s broad topic, the inquiry’s content, and how they handled an inquiry, with de-identified information available internally upon request.

Because of the uncertain nature of the COVID-19 pandemic at the time, the number of inquiries that CDC-INFO received increased from 213 255 during the year before the pandemic (January 2019, 2019, to January 19, 2020) to 941 745 during the first year of the pandemic (January 20, 2020, to January 20, 2021) (email communication, Su Yeon Bae, CDC, February 10, 2023). Many of these inquiries related to K-12 schools. We (D.K., L.C.B., N.K.C.) responded to many of these inquiries and recognized the opportunity to learn from this experience. This analysis of prior inquiries may inform public health guidance for schools before, during, and after emergencies and improve how health officials respond to public inquiries during a crisis.

Our study had 4 primary research questions:

1. How often did CDC receive inquiries about various K-12 school topics?
2. How did the frequency of K-12 school questions versus complaints vary by theme?
3. How did the frequency of K-12 school themes vary over time?
4. How did CDC-INFO agent response sources (website, prepared responses, escalation to subject matter experts, or no action needed) for K-12 school topics vary by theme?

## Methods

### Study Sample

This study is a retrospective analysis of secondary data from CDC’s CDC-INFO inquiry tracking system. This activity was reviewed by CDC, deemed not research, and conducted consistent with applicable federal law and CDC policy (accession # NCHHSTP-DASH-2/15/23-befa8). An inquiry may include 1 or more questions or complaints.

CDC-INFO statisticians compiled a report using Power BI Desktop Version: 2.109.782.0 (Microsoft Corp) containing all CDC-INFO inquiries that included the word “school” (the setting of interest) and excluded the words “invite” or “interview” (invitations for interviews or speakers were excluded as not directly related to K-12 guidance content). In this report, only inquiries assigned 1 or more of the following topics by CDC-INFO agents were included: “coronavirus”, “novel coronavirus 2019”, or “novel coronavirus 2019, China.” This report contained inquiries from January 17, 2020 (when the first inquiry meeting inclusion criteria occurred), through November 8, 2022 (when data collection for the purpose of this study ceased).

The initial inclusion dataset contained 28 838 inquiries. When sorting by inquiry identification numbers, we identified and removed 4336 duplicates, resulting in 24 502 total inquiries. We established a process that led us to a final set of inquiries to analyze for the purpose of this study.

First, we developed a data coding protocol that listed the inclusion and exclusion criteria, along with a list of proposed themes and their definitions. We created all coding using Excel (Microsoft Corp). Before conducting the analysis, we pilot-tested this protocol with a simple random sample of 100 inquiries from the 24 502 total. We independently reviewed inquiries to determine whether the inquiry was relevant for inclusion. We considered inquiries relevant if they had a clear question or complaint and were related to COVID-19 and K-12 school staff or students in the United States. During this pilot testing, if the inquiry was relevant, we independently selected 1 or more themes describing the inquiry content. We completed 3 rounds of this process, discussing discrepancies after each round of coding, combining some of the themes, and updating definitions for clarity and to improve interrater reliability. During the first 2 rounds of testing, 7 people coded inquiries. In the final round of pilot testing, we reduced the group to 4 coders to improve consistency. We did not include the 300 pilot-tested inquiries in the final analysis. The final list of 11 themes included the following: closure, general guidance, high-risk/disability (ie, students or staff at high risk of severe disease or with a disability), prevention— cleaning, prevention—distance, prevention—face masks, prevention—vaccines, quarantine and isolation, special settings, testing, and travel. The general guidance theme typically included broad questions such as “What is CDC’s guidance on COVID-19 for K-12 schools?”

Second, after completing pilot testing, we used the Cohens *κ* coefficient to analyze interrater reliability for the final round of 100 pilot inquiries on the decision to keep or exclude an inquiry and how often the coders independently selected the same theme. To ensure good interrater reliability, we assessed Cohen *κ* values between each pair of coders. We then validated agreement with all 4 coders combined by using the Gwet AC1 statistic. For both the Cohen *κ* coefficient and the Gwet AC1 statistic, we considered agreement to be very good if *κ* was 0.8 to 1 and good if *κ* was 0.6 to 0.8.^3^

For the analysis, we targeted having at least 30 inquiries for each of the 11 themes, which led to a minimum target sample size of 330 inquiries. We increased the sample size by approximately 12 times (to 4000 inquiries) because a large number of inquiries would not be relevant, sufficient cell sizes would also be needed for the period and disposition analyses, and the number of relevant inquiries per theme was unevenly distributed. After completing pilot testing and ensuring sufficient interrater reliability, the 4 authors (D.K., L.C.B., N.K.C., W.R.) who coded inquiries during pilot testing were each assigned a random sample of 1000 inquiries from the remaining 24 202 inquiries to review for relevancy and to code with 1 or more themes. We regularly met throughout the coding process to discuss how to assign complex inquiries. Of the 4000 inquiries reviewed, we identified 2180 as relevant for the analysis and coded them accordingly. The number of relevant inquiries exceeded the minimum target sample size of 330 inquiries for analysis and ensured that even themes with fewer inquiries had at least 30 inquiries. We considered the remaining 1820 inquiries irrelevant and excluded them from analysis. For example, we excluded inquiries that were about non–K-12 schools (eg, preschools, colleges), those asking for guidance for schools outside the United States, or any inquiries without a clear question or complaint.

Finally, for all inquiries that included multiple themes, we duplicated entries so that the identification number was represented once per relevant theme to ensure that all themes would be captured in the analysis. Of the 2180 inquiries retained as relevant, 251 included multiple themes, resulting in 2439 themes coded. We noted whether each inquiry included a complaint or a question and excluded inquiries that consisted only of a comment.

### Statistical Analyses

To understand how the frequency of themes varied over time, we split the 1027 days of data collected into 5 sections of 204 to 205 days. We chose 5 sections because each section captured multiple key events in the COVID-19 pandemic^1^ (Table 1) and school guidance updates and were also feasible for analysis. These sections included time 1 (T1): January 17–August 8, 2020; time 2 (T2): August 9, 2020– March 1, 2021; time 3 (T3): March 2–September 23, 2021; time 4 (T4): September 24, 2021–April 17, 2022; and time 5 (T5): April 18–November 8, 2022.

**Table 1.**
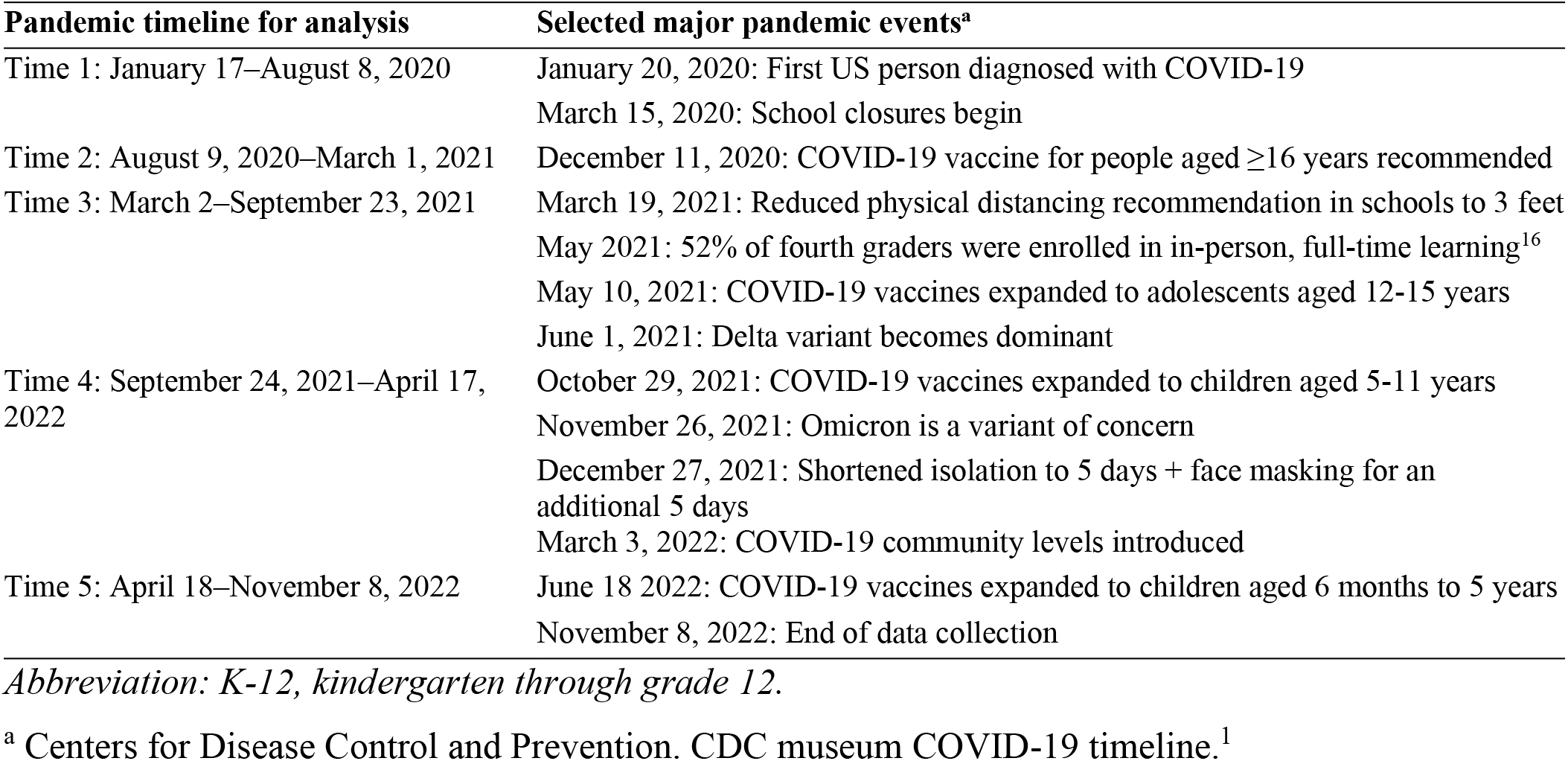
Time frames for analysis and alignment with COVID-19 pandemic events affecting K-12 schools, United States, 2020-2022.

CDC-INFO agents assign each inquiry a disposition, which represents how they responded to the inquirer. To understand how CDC-INFO response sources varied by theme, we combined the 12 disposition categories represented in this project into 4 key categories for analysis:

1. Prepared response: Existing category used when an agent shared a pre-created response to common questions, which allows them to answer questions quickly.
2. Website: Existing category used when an agent shared information directly from the CDC website in response to the inquiry.
3. Escalation: Used when an agent escalates the inquiry to subject matter experts at CDC for response. This new category included the following original disposition categories: clarification (email only), escalation, program referral, program response, and transferred.
4. No CDC action: Used when an agent determines no further action is needed. This new category included the following original disposition categories: agent terminated, call dropped, external referral, for your information, and publication request. Examples include questions or complaints on topics outside CDC’s role, such as decisions on school closures, letters allowing students to end their quarantine or isolation period, and requests to report schools for not following CDC guidelines.

We conducted Pearson χ^2^ goodness-of-fit tests to compare differences between frequencies of categorical variables, with *P* <.05 considered significant. We used SAS version 9.4 (SAS Institute Inc) to conduct all analyses.

## Results

### Interrater Reliability

The Cohen *κ* for interrater reliability between coders for whether to keep or exclude an inquiry ranged from 0.69 to 0.76 when compared with 1 other rater, indicating a substantial degree of agreement. The combined interrater reliability of all 4 coders together resulted in a Cohen *κ* of 0.73, which is considered good agreement. The *κ* statistic for agreement on the primary theme ranged from 0.59 to 0.74, with a combined *κ* of 0.67, which is considered good agreement. The Gwet AC1 statistic was 0.73, which is considered good agreement and sufficient to move from pilot to larger-scale analyses.

### Theme Frequencies

We coded a total of 2439 themes. Quarantine and isolation was the most common theme (38.7% of inquiries), followed by general guidance (20.5%), and prevention—face masks (12.1%) (Figure 1).

**Figure 1.**
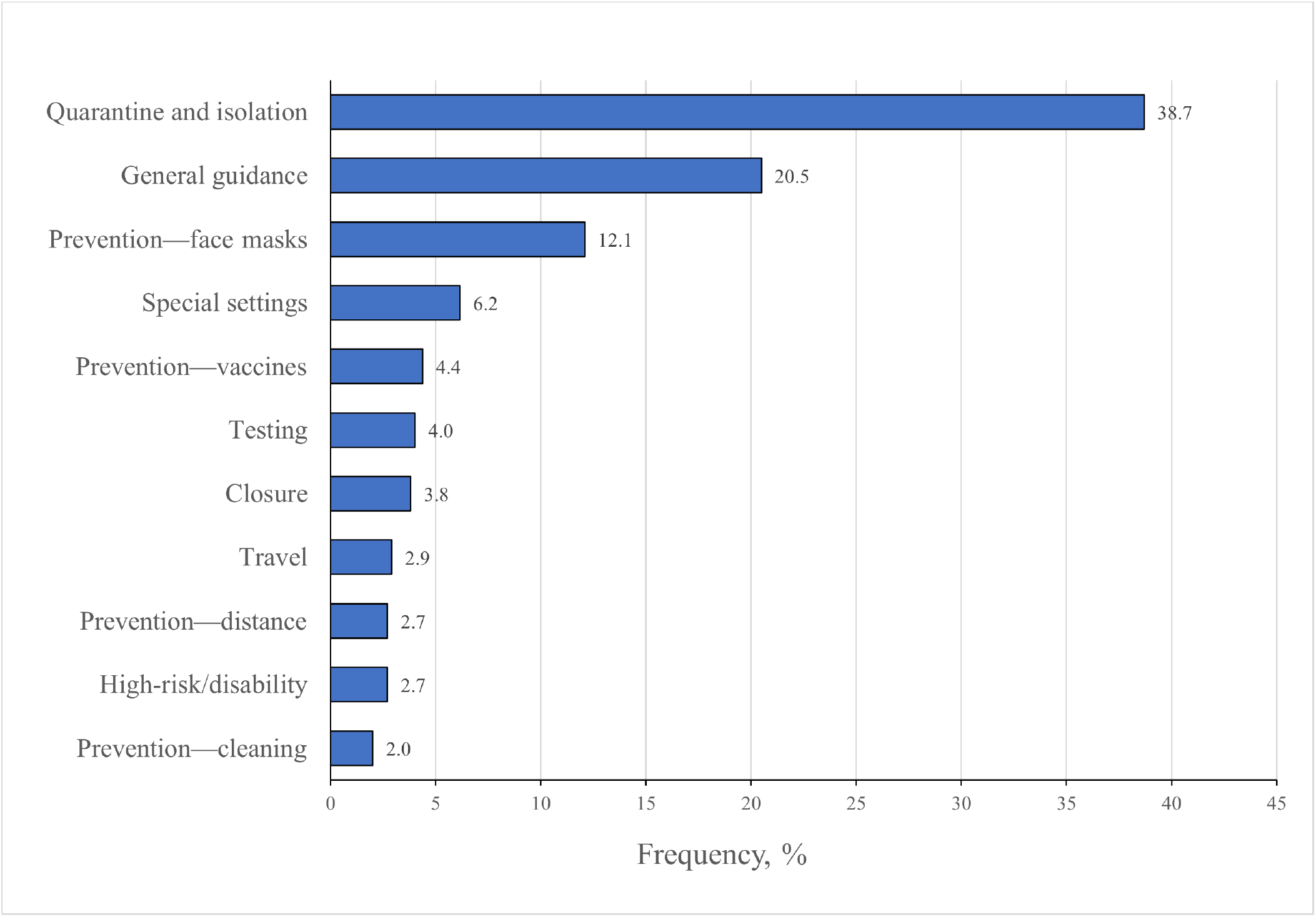
Frequency of themes (N = 2439) for K-12–related inquiries received by the Centers for Disease Control and Prevention during the COVID-19 pandemic, United States, January 17, 2020, to November 8, 2022. Abbreviation: K-12, kindergarten through grade 12

### Questions Versus Complaints

Most inquiries received were questions (74.2%) rather than complaints (25.8%). For all themes except general guidance, we found a significant difference (*P* < .05) between the proportion of questions and the proportion of complaints. Inquiry themes with a significantly higher proportion of questions than complaints included high-risk/disability (81.8% vs 18.2%), prevention—cleaning (87.8% vs 12.2%), and prevention—distance (74.2% vs 25.8%) (Figure 2). The only inquiry theme with a significantly lower proportion of questions than complaints was closure (30.1% vs 69.9%).

**Figure 2.**
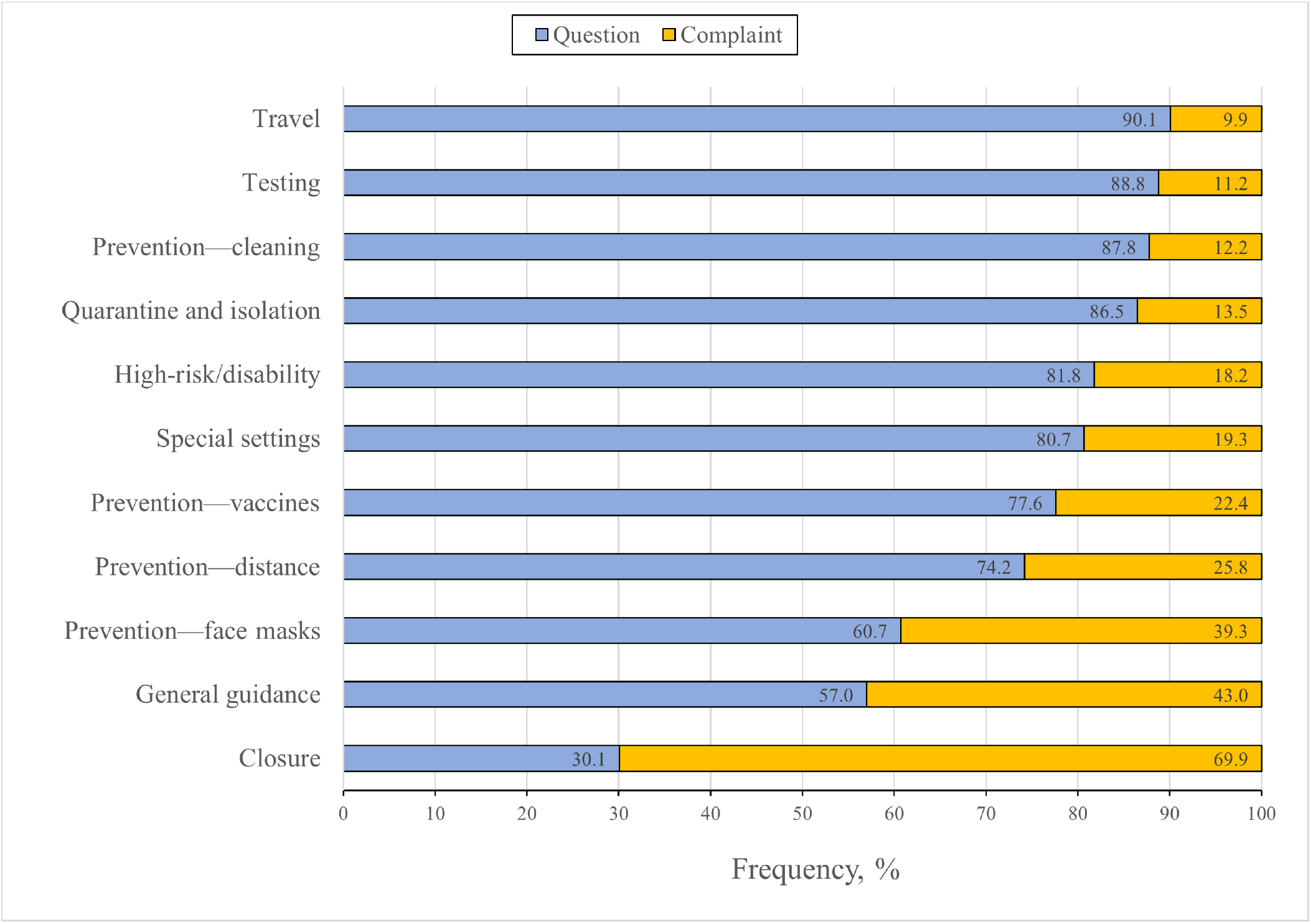
Frequency of questions versus complaints (N = 2439) by theme for K-12–related inquiries received by the Centers for Disease Control and Prevention during the COVID-19 pandemic, United States, January 17, 2020, to November 8, 2022. Abbreviation: K-12, kindergarten through grade 12. Pearson χ^2^ goodness-of-fit tests were conducted to compare differences between frequencies of categorical variables, with *P* <.05 considered significant. Only the general guidance theme was not significant.

### Frequency Over Time

Because of a low frequency of inquiries (n = 112) during T5, we removed this period from the χ^2^ comparison. The proportion of inquiries differed significantly across the remaining 4 periods for 5 of the 11 themes: closure (*P* = .01), general guidance (*P* < .001), prevention—face masks (*P* < .001), quarantine and isolation (*P* < .001), and travel (*P* = .02) (Table 2). Inquiries about closure (49 of 471; 10.4%), general guidance (192 of 471; 4.8%), and travel (39 of 471; 8.3%) were most common during T1, while inquiries about prevention—face masks (177 of 688; 25.7%) were most common during T3, and inquiries about quarantine and isolation were most common during T4 (339 of 549; 23.6%).

**Table 2.** Significance of theme frequency for K-12–related inquiries received by CDC during the COVID-19 pandemic, by period, United States^a^.

| Theme | Total frequency (T1–T4) | Time 1 (T1: January 17–August 8, 2020), frequency (%) of T1 | Time 2 (T2: August 9, 2020–March 1, 2021), frequency (%) of T2 | Time 3 (T3: March 2–September 23, 2021), frequency (%) of T3 | Time 4 (T4: September 24, 2021–April 17, 2022), frequency (%) of T4 | $\chi^2$ value (df) [*P* value]<sup>b</sup> |
| --- | --- | --- | --- | --- | --- | --- |
| Closure | 92 | 49 (10.4) | 21 (3.4) | 18 (2.6) | 4 (0.7) | 11 (3) [.01] |
| General guidance | 483 | 192 (40.8) | 143 (23.1) | 96 (14.0) | 52 (9.5) | 27 (3) [<.001] |
| High-risk/disability | 63 | 19 (4.0) | 22 (3.6) | 15 (2.2) | 7 (1.3) | 2 (3) [.48] |
| Prevention—cleaning | 47 | 14 (3.0) | 16 (2.6) | 12 (1.7) | 5 (0.9) | 1 (3) [.75] |
| Prevention—distance | 66 | 14 (3.0) | 18 (2.9) | 32 (4.7) | 2 (0.4) | 4 (3) [.29] |
| Prevention—face masks | 290 | 20 (4.2) | 47 (7.6) | 177 (25.7) | 46 (8.4) | 25 (3) [<.001] |
| Prevention—vaccines | 94 | 0<sup>c</sup> | 31 (5.0) | 45 (6.5) | 18 (3.3) | 2 (2)<sup>c</sup> [.45] |
| Quarantine and isolation | 877 | 75 (15.9) | 254 (41.0) | 209 (30.4) | 339 (61.7) | 30 (3) [<.001] |
| Special settings | 149 | 34 (7.2) | 42 (6.8) | 45 (6.5) | 28 (5.1) | 0 (3) [.93] |
| Testing | 96 | 15 (3.2) | 15 (2.4) | 31 (4.5) | 35 (6.4) | 3 (3) [.48] |
| Travel | 70 | 39 (8.3) | 10 (1.6) | 8 (1.2) | 13 (2.4) | 10 (3) [.02] |
| Total | 2327 | 471 (20.2) | 619 (26.6) | 688 (29.6) | 549 (23.6) |  |

Abbreviations: CDC, Centers for Disease Control and Prevention; K-12, kindergarten through grade 12.
<sup>a</sup> Because of a low frequency of inquiries (n = 112) during T5 (April 18–November 8, 2022), this period was removed from the $\chi^2$ comparison.
<sup>b</sup> Pearson $\chi^2$ goodness-of-fit tests were conducted to compare differences between frequencies of categorical variables, with *P* < .05 considered significant.
<sup>c</sup> No inquiries were expected for T1 related to vaccines because the COVID-19 vaccine did not become available until December 11, 2020, with the first vaccines only available to those aged ≥16 years.<sup>1</sup>

### Themes and Dispositions

For all themes except travel, we found significant differences in how CDC-INFO agents responded to inquiries. Agents most often used prepared responses to answer inquiries (986 of 2439; 40.4%), followed by escalation (497 of 2439; 20.4%), and website (355 of 2439; 14.6%) (Table 3). An additional 24.6% of inquiries (601 of 2439) required no CDC action.

**Table 3.** Significance of theme frequency for K-12–related inquiries received by CDC during the COVID-19 pandemic, by how CDC-INFO agents responded to inquiries, United States, January 17, 2020, to November 8, 2022^a^.

| Theme | Total frequency, no. | Website, no. (%) | Prepared response, no. (%) | Escalation,<sup>a</sup> no. (%) | No CDC action,<sup>b</sup> no. (%) | $\chi^2$ (df) [*P* value]<sup>c</sup> |
| --- | --- | --- | --- | --- | --- | --- |
| Closure | 93 | 6 (6.5) | 20 (21.5) | 20 (21.5) | 47 (50.5) | 40 (3) [$<.001$] |
| General guidance | 500 | 48 (9.6) | 178 (35.6) | 115 (23.0) | 159 (31.8) | 16 (3) [.001] |
| High-risk/disability | 66 | 9 (13.6) | 20 (30.3) | 24 (36.4) | 13 (19.7) | 12 (3) [.01] |
| Prevention—cleaning | 49 | 8 (16.3) | 14 (28.6) | 21 (42.9) | 6 (12.2) | 24 (3) [$<.001$] |
| Prevention—distance | 66 | 9 (13.6) | 26 (39.4) | 16 (24.2) | 15 (22.7) | 13 (3) [.005] |
| Prevention—face masks | 295 | 32 (10.8) | 92 (31.2) | 75 (25.4) | 96 (32.5) | 12 (3) [.01] |
| Prevention—vaccines | 107 | 12 (11.2) | 45 (42.1) | 22 (20.6) | 28 (26.2) | 20 (3) [$<.001$] |
| Quarantine and isolation | 944 | 184 (19.5) | 470 (49.8) | 117 (12.4) | 173 (18.3) | 35 (3) [$<.001$] |
| Special settings | 150 | 21 (14.0) | 53 (35.3) | 45 (30.0) | 31 (20.7) | 11 (3) [.01] |
| Testing | 98 | 12 (12.2) | 43 (43.9) | 24 (24.5) | 19 (19.4) | 23 (3) [$<.001$] |
| Travel | 71 | 14 (19.7) | 25 (35.2) | 18 (25.4) | 14 (19.7) | 6 (3) [.11] |
| Total | 2439 | 355 (14.6) | 986 (40.4) | 497 (20.4) | 601 (24.6) | — |

Abbreviations: CDC, Centers for Disease Control and Prevention; K-12, kindergarten through grade 12.
<sup>a</sup> Includes the following disposition categories: clarification (email only), escalation, program referral, program response, and transferred.
<sup>b</sup> Includes the following disposition categories: agent terminated, call dropped, external referral, for your information, and publication request.
<sup>c</sup> Pearson $\chi^2$ goodness-of-fit tests were conducted to compare differences between frequencies of categorical variables, with $P < .05$ considered significant.

Compared with other methods CDC-INFO agents used to respond to inquiries, closure and prevention—face masks received significantly higher proportions of inquiries requiring no CDC action (50.5% and 32.5%, respectively), while general guidance, prevention—distance, prevention—vaccines, quarantine and isolation, special settings, and testing had significantly higher proportions of inquiries answered with prepared responses (35.6%, 39.4%, 42.1%, 49.8%, 35.3%, 43.9%, respectively) (Table 3).

Inquiries related to high-risk/disability, prevention—cleaning, and special settings resulted in a greater proportion of escalations to subject matter experts compared with other response sources (36.4%, 42.9%, and 30.0%, respectively) (Table 3).

## Discussion

The COVID-19 pandemic highlighted the need for rapid sharing of public health guidance in an ever-changing environment. This need for rapid sharing was especially critical for K-12 schools, where recommendations for staff and students often included school closures and quarantine and isolation periods.^4^ Public uncertainty^5^ may have resulted in an increased number of inquiries to CDC related to public health guidance. Inquiry themes also may have changed based on which topics were included in news coverage or social media posts. During the period reviewed, CDC received 24 502 inquiries related to K-12 schools and COVID-19. This number is much higher than the number of inquiries received for other populations in other emergencies; for example, the 4661 inquiries related to 2009 H1N1 and pregnant, postpartum, or breastfeeding women in a prior study of H1N1 inquiries^6^ and the 108 inquiries from an Ebola and maternal health study.^7^ This increase in inquiries may be a result of the large proportion of the population who are K-12 students, their families, or staff members; the long time frame and widespread nature of the COVID-19 pandemic; and the effects of school closures and quarantine and isolation, such as disruptions to parents’ ability to work and the need for families to navigate increased caregiving responsibilities and conflict.^4,8^

CDC received many questions about COVID-19 quarantine and isolation related to K-12 settings, which is understandable because it had a large effect on individuals’ daily lives.^8^ Interestingly, the frequency of inquiries about quarantine and isolation was greater than the frequency of inquiries related to prevention in the K-12 setting, perhaps because of the aforementioned effect on families.

Similarly, a study focusing on maternal health inquiries during the 2014-2015 Ebola response found that infection control was the most common inquiry topic,^7^ and a study focusing on hepatitis C inquiries found that transmission was the second most common inquiry topic.^9^ However, during the H1N1 pandemic, inquiries related to pregnant women were primarily related to vaccines (69%), with fewer inquiries related to infection control (13%).^6^ The later accessibility of COVID-19 vaccines for children than for adults, as well as our study’s focus on the school setting, likely captured fewer vaccine-related inquiries than the study of pregnant women during H1N1. Although travel was included in only 2.9% of inquiries related to COVID-19 and K-12 schools, it was one of the most frequent inquiry topics (42%) during the 2015 Zika outbreak, whereas transmission constituted only 9% of inquiries.^10^ The low frequency of travel-related inquiries in this study compared with the study during Zika may be a result of the K-12 school–specific focus rather than the overall population, because Zika was primarily being transmitted outside the United States, or the primary transmission of Zika through mosquitoes rather than close person-to-person contact,^11^ which may have affected risk perceptions.

The higher frequency of questions than complaints overall highlights the public’s need for information that is easy to access, understand, and apply, especially during times of uncertainty, such as during a novel pandemic.^12^ The topic of closures received many more complaints than questions, likely because of frustration at the large effect it had on families and staff, many of whom faced new childcare challenges.^13^ Interestingly, although also impactful, quarantine and isolation received many more questions than complaints, perhaps because of the relatively shorter effect of quarantine and isolation (often <14 d vs semester-long closures).^14^

The number of inquiries gradually increased from T1, to T2, to T3, slowly dropped during T4, and then declined sharply in T5. The gradual increase may provide insight about what public health professionals can expect in future responses. Understandably, most frequencies in themes decreased over time as initial questions were answered and more information was available online. However, questions about prevention—face masks increased significantly during T3, likely because of changes in social distancing recommendations from 6 feet to 3 feet in the school setting while wearing face masks,^15^ along with many students returning to in-person learning.^16^ The frequency of the quarantine and isolation theme increased in T2, decreased in T3, and then increased again in T4. This fluctuation in inquiry frequency was likely because of the alignment of the time period with the school year, along with decreases in contact tracing and changing quarantine recommendations over time.^17^

CDC-INFO agents managed inquiries several ways. The high proportion of escalations to subject matter experts related to the high-risk/disability theme was likely because students and staff at increased risk or with a disability may have unique situations^18^ that are unable to be fully addressed by general guidance. For prevention—cleaning inquiries, the high proportion of escalations may indicate a need for improved guidance on the website or through prepared response materials to respond to these inquiries. Additionally, many inquiries required no CDC action, such as those focused on school closure, which were not decisions made by CDC.^19^ The high frequency of inquiries that did not require CDC action highlights potential misunderstandings of CDC’s role during a public health emergency.

### Limitations

This study had several limitations. First, this study included only a portion of the inquiries directed toward CDC; therefore, it may not be fully generalizable to local public health agencies or be representative of the questions and complaints the public had. Second, this study did not examine the relationship between the inquiries and changes in K-12 guidance, which would require reviewing considerably more inquiries and conducting more complex analyses. Third, this study did not compare Finally, this study only focused on COVID-19. Additional studies of this population during other public health events would help to clarify how needs may change by public health threat. For example, a radiological event may raise different questions than an outbreak of influenza or a novel pathogen response.

## Conclusions

Public health officials should be prepared to respond to the changing informational needs of the public during emergency responses and to provide both general information through websites and detailed information for unique situations and settings, including K-12 schools. In future responses, public health officials should proactively be prepared to respond to questions about quarantine and isolation and to address various topics as the emergency changes over time. Public health officials should also be prepared for complaints about school closures and to dedicate subject matter experts to responding to some questions, such as those related to high-risk/disability, cleaning guidance, and special settings. Public health preparation and response can help empower individuals to take steps to protect their health and their communities. Additional research is needed on an ongoing basis to better facilitate public health officials’ ability to rapidly adapt materials for the needs of their communities, tailoring materials to specific populations and responses.

## Data Availability

All data produced in the present study are available upon reasonable request to the authors.

## Disclaimer

The findings and conclusions of this article are those of the authors and do not represent the views of the Centers for Disease Control and Prevention.

## References

1. Centers for Disease Control and Prevention. CDC museum COVID-19 timeline. Updated August 16, 2022. Accessed February 10, 2023. https://www.cdc.gov/museum/timeline/covid19.html

2. Centers for Disease Control and Prevention.CDC-INFO. Updated November 28, 2022. Accessed February 10, 2023. https://www.cdc.gov/cdc-info/index.html

3. Altman DG. Practical Statistics for Medical Research. Chapman & Hall/CRC Press; 1999.

4. Gupta S, Smith L, Diakiw A. Avoidance of COVID-19 for children and adolescents and isolation precautions. Pediatr Clin North Am. 2021;68(5):1103–1118. doi:10.1016/j.pcl.2021.05.011

5. Strydhorst NA, Landrum AR. Charting cognition: mapping public understanding of COVID-19. Public Underst Sci. 2022;31(5):534–552. doi:10.1177/09636625221078462

6. Mosby LG, Ellington SR, Forhan SE, et al. The Centers for Disease Control and Prevention’s maternal health response to 2009 H1N1 influenza. Am J Obstet Gynecol. 2011;204(6 Suppl 1): S7–S12. doi:10.1016/j.ajog.2011.02.057

7. Ellington S, Perez M, Morof D, et al. Addressing maternal health during CDC’s Ebola response in the United States. J Womens Health (Larchmt). 2017;26(11):1141–1145. doi:10.1089/jwh.2017.6719

8. Gayatri M, Puspitasari MD. The impact of COVID-19 pandemic on family well-being: a literature review. Fam J Alex Va. 2023;31(4):606–613. doi:10.1177/10664807221131006

9. Jorgensen CM, Lewis CA, Liu J. An analysis of hepatitis C virus–related public inquiries from health professionals: 2009-2010. Clin Infect Dis. 2012;55(Suppl 1):S54-S57. doi:10.1093/cid/cis369

10. Sell TK, Watson C, Meyer D, et al. Zika inquiries made to the CDC-INFO system, December 2015–September 2017. Emerg Infect Dis. 2020;26(5):1022–1024. doi:10.3201/eid2605.181694

11. Centers for Disease Control and Prevention. Zika cases in the United States. Updated May 15, 2024. Accessed June 17, 2024. https://www.cdc.gov/zika/zika-cases-us/index.html

12. Eldridge CC, Hampton D, Marfell J. Communication during crisis. Nurs Manage. 2020;51(8):50–53. doi:10.1097/01.NUMA.0000688976.29383.dc

13. Whaley GL, Pfefferbaum B. Parental challenges during the COVID-19 pandemic: psychological outcomes and risk and protective factors. Curr Psychiatry Rep. 2023;25(4):165–174. doi:10.1007/s11920-023-01412-0

14. Dawson P, Worrell MC, Malone S, et al. Modifications to student quarantine policies in K-12 schools implementing multiple COVID-19 prevention strategies restores in-person education without increasing SARS-CoV-2 transmission risk, January–March 2021. PLoS One. 2022;17(10):e0266292. doi:10.1371/journal.pone.0266292

15. Jenco M. CDC changes recommended distance between students to 3 ft. while masked. AAP News. March 19, 2021. Accessed June 17, 2024. https://publications.aap.org/aapnews/news/7908/CDC-changes-recommended-distance-between-students

16. Merod A, Arundel K. As national COVID-19 emergency ends, a look back on the virus’ impact on schools. K-12 Dive. May 11, 2023. Accessed July 31, 2024. https://www.k12dive.com/news/national-emergency-ends-COVID-19-timeline/650009

17. Zou K, Hayashi M, Simon S, Eisenberg JNS. Trade-off between quarantine length and compliance to optimize COVID-19 control. Epidemiology. 2023;34(4):589–600. doi:10.1097/EDE.0000000000001619

18. Blad E. Special education during the pandemic, in charts. EducationWeek. October 17, 2022. Accessed July 31, 2024. https://www.edweek.org/teaching-learning/special-education-during-the-pandemic-in-charts/2022/10

19. Zviedrite N, Hodis JD, Jahan F, Gao H, Uzicanin A. COVID-19–associated school closures and related efforts to sustain education and subsidized meal programs, United States, February 18–June 30, 2020. PLoS One. 2021;16(9):e0248925. doi:10.1371/journal.pone.0248925

